# Large-scale seroepidemiologic surveillance of COVID-19 - Cross-sectional study in Hyogo prefecture of Japan in August, 2021

**DOI:** 10.1101/2021.09.26.21264129

**Authors:** Zhenxiao Ren, Koichi Furukawa, Mitsuhiro Nishimura, Yukiya Kurahashi, Silvia Sutandhio, Lidya Handayani Tjan, Kaito Aoki, Natsumi Hasegawa, Jun Arii, Kenichi Uto, Keiji Matsui, Itsuko Sato, Jun Saegusa, Nonoka Godai, Kohei Takeshita, Masaki Yamamoto, Tatsuya Nagashima, Yasuko Mori

## Abstract

The situation of the COVID-19 pandemic in Japan is drastically changing in the 2^nd^ year, 2021, due to the appearance of SARS-CoV-2 variants of concern and the roll-out of mass vaccination. In addition to PCR diagnosis, periodic seroepidemiologic surveillance is important to analyze the epidemic situation. In this study, we analyzed the rate of seropositivity for the SARS-CoV-2 N and S antigens in Hyogo prefecture, Japan in August 2021. Sera collected from people who received a health check-up in a clinic of the Hyogo Prefecture Health Promotion Association were subjected to analysis of reactivity to the SARS-CoV-2 N and S antigens by electrochemiluminescence immunoassay (ECLIA) and enzyme-linked immunosorbent assay (ELISA), respectively. For a total 1,000 sera, the positive rates to N and S antigens were 2.1% and 38.7%, respectively. The infectious rate estimated by serological analysis based on the presence of the anti-N antibody was 2.5-fold higher than the value reported based on PCR-based analysis, and it increased five-fold compared to the rate determined by our previous seroepidemiologic study in October, 2020. The anti-S positive rate was almost consistent with the vaccination rate in this area. The observed high anti-S antibody level in the seropositive population may indicate that the mass vaccination in Japan is being performed smoothly at this time point, although the infectious rate has also increased.

## Introduction

The COVID-19 pandemic, which first emerged in December 2019, has undergone several turning points, introducing drastic changes in its progress. One of the most important factors is the appearance of the SARS-CoV-2 variants of concern (VOCs) replacing the original variant. In Japan, the Alpha variant (B1.1.7) replaced the existing strain by around April, 2021; then, the Delta variant (B1.617.2) began spreading rapidly throughout the country from July to August, 2021 (2,16,22). The other critical event has been the launch of anti-COVID-19 vaccines. Accelerated development of several vaccine platforms was based on the components of the original SARS-CoV-2 strain as the template, and these vaccines were shown to be effective for reducing the COVID-19 outbreak (17, 20). Although the COVID-19 vaccine rollout in Japan was delayed compared to that in other countries such as Israel and United States (19), the Comirnaty (BNT162b2 mRNA, BioNTech-Pfizer, Mainz, Germany/New York, United States) vaccine (25) was first approved in Japan and administered to health-care workers starting in February 2021, and elderly persons aged ≥65 years old starting April 2021, expanding to other populations from May, 2021. The mRNA vaccines developed by Moderna (7) were also approved and have been widely used in Japan from May 2021. The number of vaccinated people is increasing rapidly in Japan, as monitored by the government system (1). On September 13th, the government announced that 50.9% of the population in Japan has completed vaccination (https://covid19.who.int/region/wpro/country/jp). These two factors, that is, propagation of SARS-CoV-2 VOCs and the progress of the vaccination, may balance each other out, making predictions of the COVID-19 situation more difficult.

In October 2020, we conducted seroepidemiologic surveillance in Hyogo prefecture, population 5.5 million, located in the southern-central region of Japan, the so-called Kansai area (11). In our 1st seroepidemiologic surveillance, we reported that 0.15% of 10,377 sera had neutralizing activity against SARS-CoV-2 infection *in vitro*. The value 0.15% was interpreted to be the true infectious rate, and it was three times higher than the value (0.05%) estimated based on the PCR diagnosis at that time point (33), probably because the serological analysis could also detect persons with histories of asymptomatic infection.

In previous COVID-19 seroepidemiologic analyses, both the Nucleocapsid (N) and Spike (S) antigens of the SARS-CoV-2 were used as markers of infection; reactivity of serum antibodies against one or two of these antigens was detected (9, 21, 27, 33). However, currently the anti-N antibody is the major marker because the vaccines used in Japan protect individuals by inducing anti-S immunities including anti S-antibodies, making it is impossible to distinguish the S antibodies due to SARS-CoV-2 infection from those induced by vaccination. Nevertheless, analysis of the anti-S antibodies is of great importance because they are the key immune components for counteracting SARS-CoV-2 infection, and can serve as a marker of an individual’s potential to be protected from the infection (29, 31).

Under the current situation where infected and vaccinated people are both prevalent in society, seroepidemiologic surveillance can be used to accurately measure the proportion of individuals who have acquired SARS-CoV-2-specific antibodies by infection, vaccination or both. Seroepidemiologic surveillance is frequently conducted to understand and predict epidemics (8, 23, 26, 28, 35).

In this study, we reported the 2^nd^ seroepidemiologic survey of COVID-19 in Hyogo prefecture from July to August 2021, which overlaps with the period of the 2020 Tokyo Olympics, delayed one year and held in Japan in the summer of 2021 (30). The anti-N antibody positivity rate revealed that the estimated infection rate was slightly higher than that reported by PCR diagnosis, whereas the anti-S antibody positive rate was consistent with the current vaccination rate in Hyogo prefecture. This report will be a milestone in the era during which the vaccine and SARS-CoV-2 VOCs are struggling for dominance.

## Materials and methods

### Samples

Serum samples were collected from people receiving health check-ups at the clinics of Hyogo Prefecture Health Promotion Association, Kobe, Japan from 19^th^ July to 6^th^ August. This retrospective observational study was explained on the website of Kobe University Hospital along with the opportunity to opt-out. Each serum was heat-treated at 56°C for 30 min and stored in 4°C until use.

### Detection of anti-N antibody by electrochemiluminescence immunoassay (ECLIA)

An electrochemiluminescence immunoassay (ECLIA) was conducted using the cobas e801 module (Roche Diagnostics, Rotkreuz, Switzerland) as in our previous study (11). The Elecsys Anti-SARS-CoV-2 assay kit (Roche Diagnostics, Rotkreuz, Switzerland) is based on a double-antigen sandwich assay, which detects antibodies against the SARS-CoV-2 nucleocapsid (N). The measurement was performed according to the manufacturer’s instructions, and samples with a cut-off index (COI) > 1.0 were diagnosed as positive.

### Expression and purification of SARS-CoV-2 spike protein

The Spike protein used for the ELISA assay was prepared by a recombinant expression system similar to that previously reported by another research group (15). Briefly, the gene sequence of the SARS-CoV-2 Spike ectodomain (amino acids 1-1213) was subcloned into a pCAGGS vector (24) with a puromycin-resistant gene. The Spike sequence used here includes following mutations: D614G, R682del, R683del, R685del, F817P, A892P, A899P, A942P, K986P, and V987P, and additional sequences: namely, an HRV-3c recognition site, T4 foldon, and a His-tag at the C-terminal side. The sequence was confirmed by the capillary electrophoresis sequencer DS3000 (Hitachi High-Tech).

Spike protein was expressed using the Expi293 expression system (Thermofisher Scientific) according to the manufacturer’s instruction. The culture supernatant was collected at four days post transfection. The His-tagged Spike protein was purified by Ni-NTA agarose (Qiagen) and size exclusion chromatography using the Superose 6 increase column and AKTA pure system (Cytiva).

### Detection of anti-Spike antibodies by enzyme-linked immunosorbent assay (ELISA)

Anti-Spike antibodies in the human sera were detected by an enzyme-linked immunosorbent assay with the purified Spike ectodomain described above. Each well of the 96-well ELISA plate (Corning) was coated with 100 ng of purified Spike protein dissolved in a carbonate buffer at 4°C overnight. After washing the plate with PBS containing 0.1% Tween 20 (PBST) to remove the antigen, PBST supplemented with 1% bovine serum albumin was added as a blocking buffer followed by incubation at 4°C for 2 hours. Individual sera samples serially diluted from 1:40 to 1:5120 in the blocking buffer were added to the antigen-coated plate, and then incubated at 37°C for 1 hour. After washing with PBST, a goat anti-human IgG with conjugated horseradish peroxidase (abcam) diluted 1:10000 with PBST was added as a secondary antibody followed by incubation at 37°C for 1 hour. After washing with PBST, 100 μl per well of ABTS solution (Roche) was added as substrate, and the plate was incubated at room temperature for 40 minutes in the dark. The reaction was stopped by adding 100 μl of 1.5% (w/v) oxalic acid dehydrate solution. The optical density at wavelength 405 nm (OD_405_) was measured using the plate leader Multiskan FC (Thermofisher Scientific). Based on a preliminary experiment using sera from healthy volunteers (average 0.14; standard deviation: 0.022, n=12), we set a cut-off value of 0.3 for the 1:40 dilution to define positivity, with the aim of avoiding false positive detection. The value of the area under the curve (AUC) was used to evaluate the anti-S antibody amount (6, 12). The AUCs were calculated for the plot of OD_405_ values according to dilution factors, and an arbitrary value of 1 was given as the width for a two-fold dilution step.

### Ethics statement

This study was approved by the ethical committees of Kobe University Graduate School of Medicine (approval codes: B2156702). Hyogo Prefecture Health Promotion Association was also granted approval under the ethical committee of Kobe University Graduate School of Medicine. Information about this retrospective observational study was published on the website of Kobe University Hospital, along with the opportunity to opt out. To validate the ELISA, sera from COVID-19 patients and healthy volunteers were used under approval of the ethical committee of Kobe University Graduate School of Medicine (approval code B200200), and written informed consent was obtained from the donors.

## Results

### Summary of the samples

We collected sera from 1,000 persons who received a health check-up at a clinic of the Hyogo Prefecture Health Promotion Association in Hyogo prefecture. A summary of the samples is shown in Table 1. The gender ratio was nearly equal: males, 58.7%; females, 41.3%. The age distribution was widely spread with a median of 48 (Table 1 and Figure 1A). The history of vaccination and infection were not included as selection or exclusion criteria; thus, no prior information was referred to in this study.

**Tables 1.**
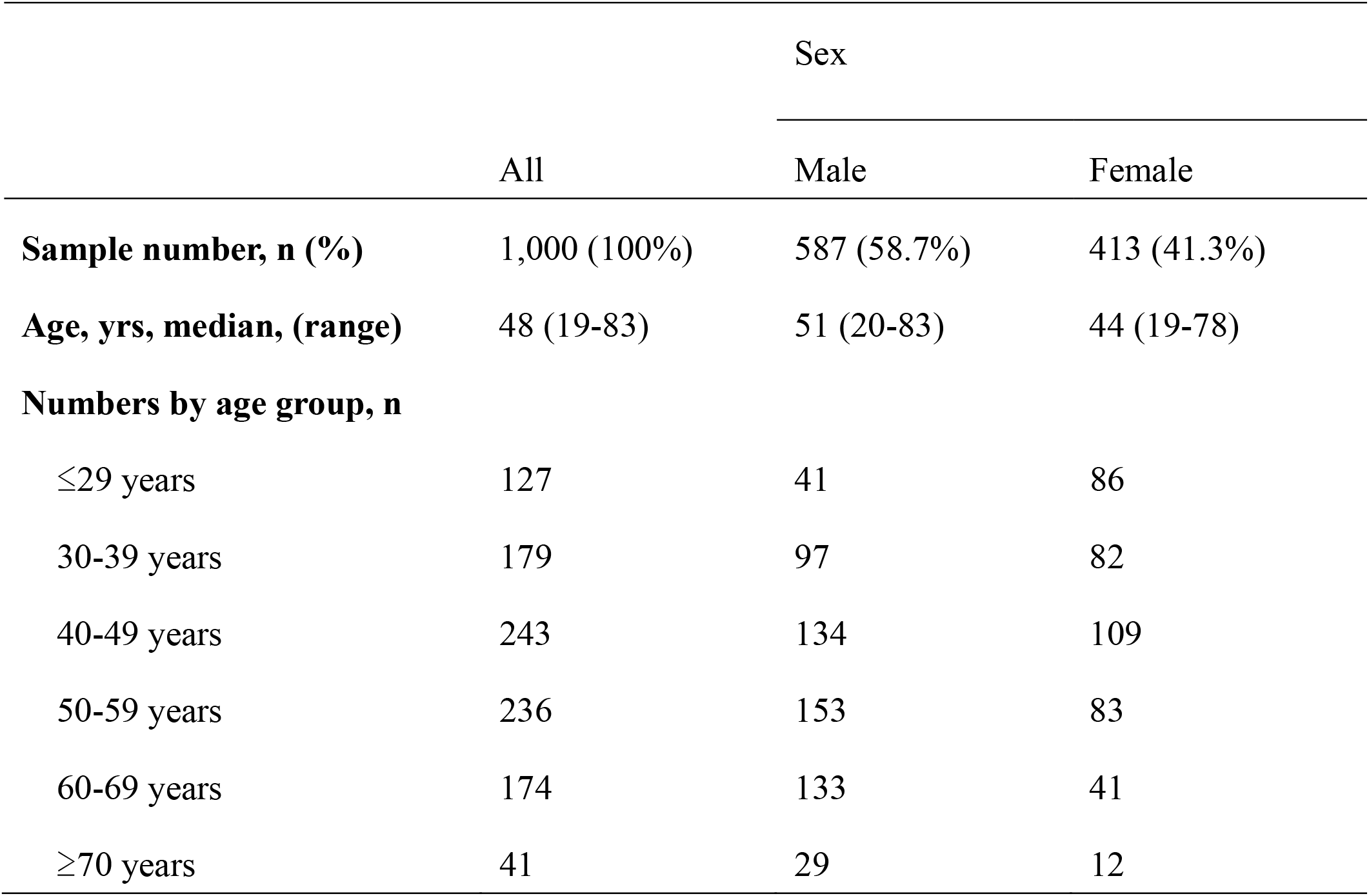
Information of the sample numbers and the age distribution.

**Figure 1.**
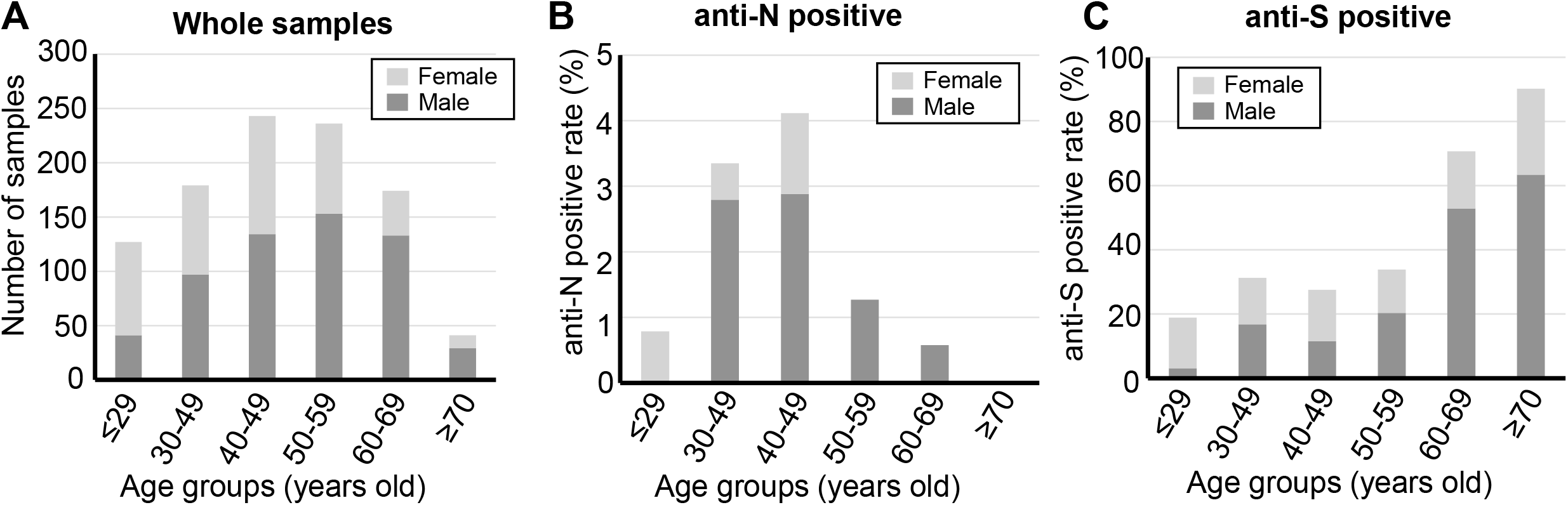
Age distribution of the samples and seropositive rate. (A) Age distribution of the volunteers who provided the sera. Subjects were divided into six age groups as follows: 29 years or less (≤29 years), 30-39 years, 40-49 years, 50-59 years, 60-69 years, 70 years or more (≥70 yrs). The anti-N positive rate (B) and anti-S positive rate (C) were calculated by age groups. The bars were painted according to the genders. For the anti-N, the measurement was performed once for all, and confirmed by a repeating measurement for the anti-N positive samples and anti-N negative samples with relatively high values (>0.25). For anti-S, all the positive samples were measured twice, and samples for which consistent results were obtained were counted as positive.

### Reactivity of sera against the SARS-CoV-2 N and S antigens

The SARS-CoV-2 anti-N antibodies were detected by the electrochemiluminescence immunoassay (ECLIA) method (10, 11), and 21 of the 1,000 samples (2.1%) were deemed positive given the cut-off of 1.0 (Table 2). The age distribution of positive cases is shown in Figure 1B and Table 2. The positive rate for the age groups 30-39 yrs and 40-49 yrs were especially high, at 3.4% (6/179) and 4.1% (10/243), respectively. The positive rates for males and females were 2.7% (16/587) and 1.2% (5/413), respectively.

**Tables 2.**
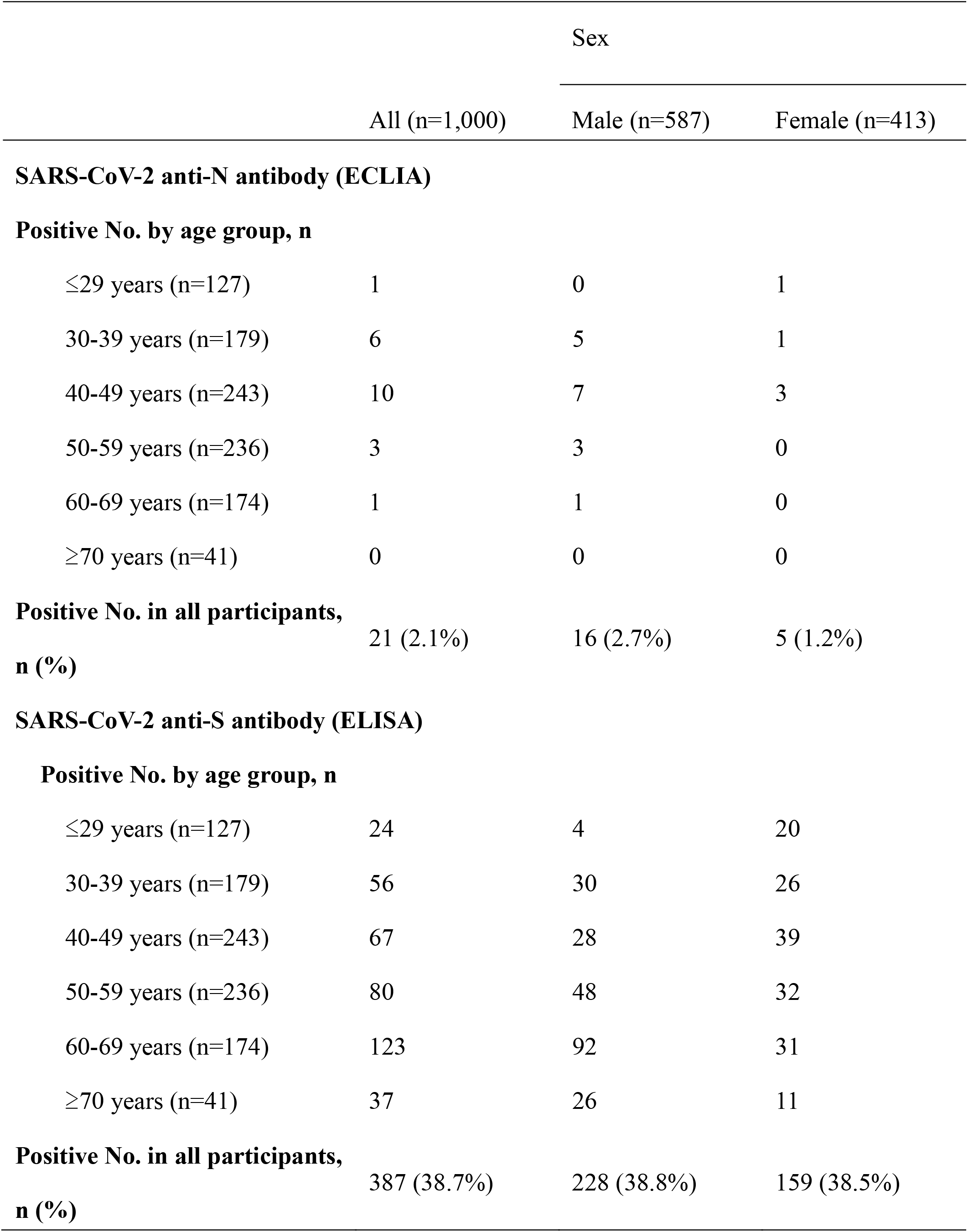
Numbers of anti-N and anti-S positive samples by age groups.

The anti-S antibodies were detected by the enzyme-linked immunosorbent assay (ELISA) method using a purified SARS-CoV-2 Spike protein. As a result, 38.7% (387/1,000) were positive given the cut-off of 0.3 (See the Materials and Methods). The age distribution of anti-S positive cases is shown in Figure 1C and Table 2. The positive rate for those in their 60s and 70s showed an apparently high rate, 70.7% (123/174) and 90.2% (37/41), respectively. On the other hand, the anti-S positive rates for younger age groups were relatively low; 18.9% (24/127), 31.3% (56/179), 27.6% (67/243), and 33.9% (80/236), for the age groups ≤29 y, 30-39 y, 40-49 y, and 50-59 y, respectively.

The correlation of measured scores for anti-N and anti-S antibodies was analyzed in N-positive cases and is shown in Figure 2. The Spearman’s correlation factor was 0.42, indicating little correlation if any, probably due to the mixed effect of infection and vaccination for the value of the anti-S antibody. As expected, all the anti-N positive sera also were anti-S positive with one exception for a serum in which the N antibody detection value was also relatively low, probably due to a weak immune response

**Figure 2.**
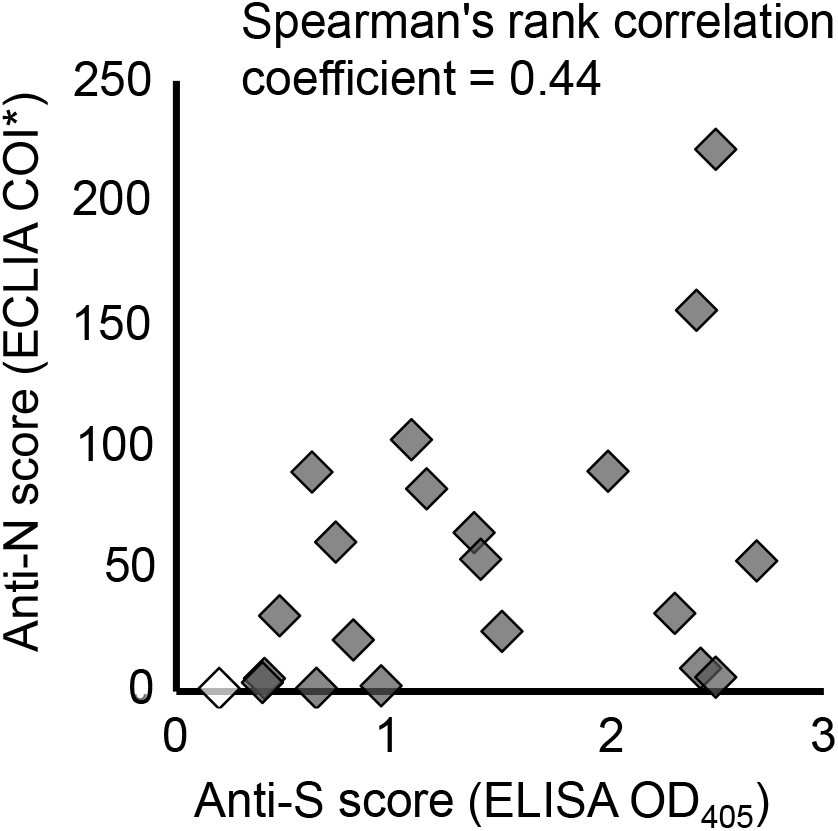
Correlation between the observed scores of anti-N and anti-S analysis. For the anti-N analysis, the cut of index value (COI) recorded by the device was used. For the anti-S analysis, the observed OD_405_ value of the ELISA was used.

### Distribution of anti-S antibody amount in sera

We further analyzed the distribution of the anti-S positive reactivity of the sera because it represents the amount of anti-S antibodies related to the neutralizing antibodies against SARS-CoV-2. The anti-S positive sera were further evaluated by serial dilution. Representative curves are shown in Figure 3A. The area under the curve (AUC), which indicates the amount of anti-S antibodies, was calculated for the 387 anti-S positive samples, and the distribution is plotted in Figure 3B. The median was 13.75, and the higher and lower quartile of the values were 17.45 and 8.79, respectively. In the same analysis, the AUC values for the serum of COVID-19 patient and that of a healthy control were 11.56 and 1.66, respectively. Thus, most of the anti-S-positive sera contained comparable amounts of anti-S antibodies to that of an actual COVID-19 patient. The anti-S AUC values for the anti-N positive sera were plotted in Figure 3C by extracting from the data of Figure 3B. The values are widely distributed within the range of whole AUC values, consistent with the result of Figure 2.

**Figure 3.**
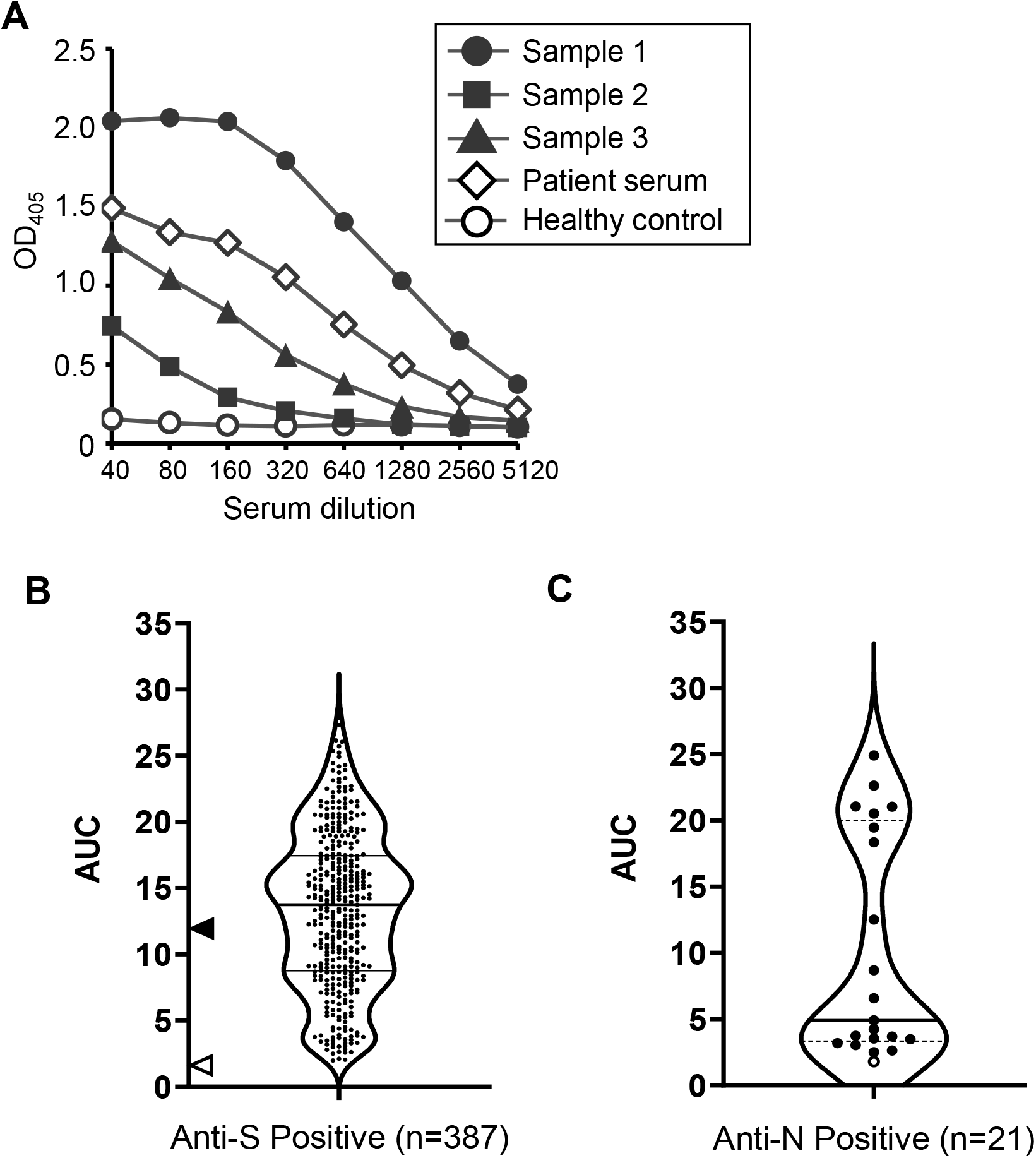
Qualitative analysis of the anti-S antibody amount. (A) Three representative curves plotting the observed OD_405_ values along the serial serum dilution factors are shown. The serum from a COVID-19 patient and that from a healthy donor were used as positive and negative controls, respectively. The areas under the curve were calculated from the individual curves and used as a reference of the anti-S amount. The calculated AUCs for the curves shown in (A) were as follows: Sample 1, 20.35; Sample 2, 3.59; Sample 3, 7.87; patient serum, 11.56; and healthy control: 1.66. (B) The distribution of AUC values from the anti-S ELISA ELISA curves are shown as a violin plot. The thick horizontal line stands for the median, 13.75, and the thin horizontal lines indicated the lower and higher quartile values, 8.79 and 17.45, respectively. The values for the patient sera and sera of healthy controls are indicated as filled and open arrowheads, respectively. (C) Distribution of anti-S AUC values for the anti-N positive sera. The exceptional sample determined as anti-S negative is shown as an open circle, and has the lowest value, as expected.

## Discussion

Seroepidemiologic surveillance is a powerful approach to understanding the spread of infectious diseases in combination with other methods such as PCR diagnosis and antigen tests. Our surveillance in August 2021 revealed that the anti-N-positive rate, which represents the SARS-CoV-2 infectious rate, was 2.1%. At the end of the surveillance, the reported infectious rate based on PCR diagnosis was 0.85% and 0.80% in the Hyogo prefecture and in Japan, respectively (33), indicating approximately a 2.5-fold difference compared with the result in this study. The difference is accounted for by the existence of asymptomatically or mildly infected individuals whose cases may not have been detected because they never underwent testing and PCR analysis. Indeed, a substantial portion of SARS-CoV-2 infected individuals are asymptomatic (4), leading to the inaccuracy of surveillance rates determined solely on PCR diagnosis. Although it is difficult to extrapolate our data directly to the whole population, they suggest that the infection is more widespread in Japan than the current PCR test results suggest and indicate a need for more systematic testing. Nonetheless, the difference between the seroepidemiologic analysis and the PCR diagnosis was not overly large; i.e., the two sets of data are mutually validating.

The infectious rate of 2.1% revealed in this study in August 2021 is 14-fold higher than the rate 0.15% we reported in our 1st seroepidemiologic analysis in October 2020 (11). The infectious rate of the 1st surveillance was based on the neutralizing antibody titer; however, vaccination has made it difficult to analyze the relationship between the infection and the neutralizing antibody titer of the sera. In contrast to the anti-S antibody, the anti-N antibody is expected to be induced only by the SARS-CoV-2 infection as long as only S-targeted vaccines are used (13, 32). The anti-N analysis by the ECLIA method of the 1st surveillance for samples collected at the same clinic of the Hyogo Prefecture Health Promotion Association was 0.4% (4/1,000) (11), indicating a 5-fold increase at the same area in these 10 months. This result was not surprising considering the fact that after the 1^st^ surveillance in October 2020, Japan including Hyogo prefecture experienced two additional waves of COVID-19 infection around January 2021 and May 2021; currently, we face the so-called fifth wave, which started in July, 2021 (3, 33).

As of August, 2021 vaccination in Japan with mRNA vaccines that induce anti-S-based immunity has been proceeding. As reported by our previous research and others, the vaccination actually induces neutralizing antibody in sera (14, 34), (Furukawa et al., manuscript submitted). According to the Japanese government system, the vaccination rate in the Hyogo prefecture as of August 6^th^, 2021 was 32.79% and 42.05% for single- or two-dose vaccinations, respectively (1). The revealed anti-S positive rate was 38.7% in this study; this result well represents the current progress in vaccination in Hyogo prefecture, Japan.

The observed high anti-S positive rate for the age groups 60-69 yrs and ≥70 yrs is explained by the priority vaccination for elderly people (more than 65 years old) in Japan (Figure 1C). The high anti-S positive rate might be related to the relatively low infection rate in these age groups (Figure 1B). However, the data for the age groups 50-59 yrs and <29 yrsr, for which both the anti-S and ant-N positive rates were relatively low, indicated that the low infectious rate could not be simply explained by the high anti-S positive rate of the age group. To understand the relatively high infectious rate for the age groups 30-39 yrs and 40-49 yrs shown in our data (Figure 1B), social reasons such as work commitments and social activities may need to be taken into consideration.

The relationship between the anti-S positive and the anti-N positive rate has become unclear because of the effect of vaccination. Actually, we could not find a correlation between the anti-N and the anti-S scores (Figure 2). Although all anti-N-positive sera were also positive for the anti-S antibody, with one exception, some of the sera showed high anti-S values with relatively low anti-N scores (Figure 2). Breakthrough infection for vaccinees may induce anti-N antibodies; however, the titers of the antibodies may be expected to be decreased owing to the prophylactic immunity induced by vaccination, preventing infection and viral replication. Actually, Allen et al. has reported that anti-N antibodies were hardly detected by the ECLIA methods we used in this study in cases of breakthrough infection (5). The cut-off value for the anti-N assays should be reconsidered in future seroepidemiologic studies in light of the high vaccination rate that is expected soon in Japan.

The COVID-19 situations in Japan and all over the world are rapidly changing day by day. This study was conducted during the build-up of social immunity by vaccination and the rapid spread of the SARS-CoV-2 Delta strain in Japan. It will become an important reference for future studies including our third epidemiological surveillance, which we anticipate will show a drastically altered situation of COVID-19.

## Data Availability

There is no available data referred to the manuscript.

## Acknowledgement

We thank Kazuro Sugimura MD, PhD (Superintendent, Hyogo Prefectural Hospital Agency and Professor, Kobe University) for his full support to promote this study. We express our sincere gratitude for cooperation and participation of staffs of Hyogo Prefecture Health Promotion Association. We also thank researchers of Division of Respiratory Medicine, Department of Internal Medicine, Kobe University Graduate School of Medicine and Hyogo Prefectural Kakogawa Medical Center.

## Abbreviations

COVID-19: Coronavirus disease 2019
SARS-CoV-2: Severe acute respiratory syndrome coronavirus 2
ECLIA: Electrochemiluminescence immunoassay
ELISA: Enzyme-linked inmmunosorbent assay
N: Nucleocapsid
S: Spike
PBS: phosphate buffered saline
COI: cut-off index
ABTS: 2,2′-azino-bis(3-ethylbenzothiazoline-6-sulfonic acid

